# Development and external validation of a prognostic multivariable model on admission for hospitalized patients with COVID-19

**DOI:** 10.1101/2020.03.28.20045997

**Authors:** Jianfeng Xie, Daniel Hungerford, Hui Chen, Simon T Abrams, Shusheng Li, Guozheng Wang, Yishan Wang, Hanyujie Kang, Laura Bonnett, Ruiqiang Zheng, Xuyan Li, Zhaohui Tong, Bin Du, Haibo Qiu, Cheng-Hock Toh

## Abstract

**Background:** COVID-19 pandemic has developed rapidly and the ability to stratify the most vulnerable patients is vital. However, routinely used severity scoring systems are often low on diagnosis, even in non-survivors. Therefore, clinical prediction models for mortality are urgently required.

**Methods:** We developed and internally validated a multivariable logistic regression model to predict inpatient mortality in COVID-19 positive patients using data collected retrospectively from Tongji Hospital, Wuhan (299 patients). External validation was conducted using a retrospective cohort from Jinyintan Hospital, Wuhan (145 patients). Nine variables commonly measured in these acute settings were considered for model development, including age, biomarkers and comorbidities. Backwards stepwise selection and bootstrap resampling were used for model development and internal validation. We assessed discrimination via the C statistic, and calibration using calibration-in-the-large, calibration slopes and plots.

**Findings:** The final model included age, lymphocyte count, lactate dehydrogenase and SpO_2_ as independent predictors of mortality. Discrimination of the model was excellent in both internal (c=0·89) and external (c=0·98) validation. Internal calibration was excellent (calibration slope=1). External validation showed some over-prediction of risk in low-risk individuals and under-prediction of risk in high-risk individuals prior to recalibration. Recalibration of the intercept and slope led to excellent performance of the model in independent data.

**Interpretation:** COVID-19 is a new disease and behaves differently from common critical illnesses. This study provides a new prediction model to identify patients with lethal COVID-19. Its practical reliance on commonly available parameters should improve usage of limited healthcare resources and patient survival rate.

**Funding:** This study was supported by following funding: Key Research and Development Plan of Jiangsu Province (BE2018743 and BE2019749), National Institute for Health Research (NIHR) (PDF-2018-11-ST2-006), British Heart Foundation (BHF) (PG/16/65/32313) and Liverpool University Hospitals NHS Foundation Trust in UK.

**Research in context:** *Evidence before this study:* Since the outbreak of COVID-19, there has been a pressing need for development of a prognostic tool that is easy for clinicians to use. Recently, a Lancet publication showed that in a cohort of 191 patients with COVID-19, age, SOFA score and D-dimer measurements were associated with mortality. No other publication involving prognostic factors or models has been identified to date.

*Added value of this study:* In our cohorts of 444 patients from two hospitals, SOFA scores were low in the majority of patients on admission. The relevance of D-dimer could not be verified, as it is not included in routine laboratory tests. In this study, we have established a multivariable clinical prediction model using a development cohort of 299 patients from one hospital. After backwards selection, four variables, including age, lymphocyte count, lactate dehydrogenase and SpO_2_ remained in the model to predict mortality. This has been validated internally and externally with a cohort of 145 patients from a different hospital. Discrimination of the model was excellent in both internal (c=0·89) and external (c=0·98) validation. Calibration plots showed excellent agreement between predicted and observed probabilities of mortality after recalibration of the model to account for underlying differences in the risk profile of the datasets. This demonstrated that the model is able to make reliable predictions in patients from different hospitals. In addition, these variables agree with pathological mechanisms and the model is easy to use in all types of clinical settings.

*Implication of all the available evidence:* After further external validation in different countries the model will enable better risk stratification and more targeted management of patients with COVID-19. With the nomogram, this model that is based on readily available parameters can help clinicians to stratify COVID-19 patients on diagnosis to use limited healthcare resources effectively and improve patient outcome.

## Introduction

Since the outbreak of coronavirus disease 2019 (COVID-19) in December 2019 in China, there have been over 200,000 confirmed cases, with 10-20% developing severe COVID-19 and approximately 5% requiring intensive care^1,2^. Although the number of daily cases has significantly reduced in China because of intensive control procedures, the numbers in the rest of world are continuing to increase. More than 10,000 deaths have been reported but the estimated mortality rate (3%) is much lower than Severe Acute Respiratory Syndrome (SARS) in 2003 (9·6%, 774 died of 8096 infected) and Middle East Respiratory Syndrome (MERS) in 2012 (34·4%, 858 died of 2494 infected)^3^. Although the vast majority of cases of COVID-19 are not life-threatening, stratification of these patients becomes increasingly important as large populations are expected to be infected globally.

The deaths and morbidity from COVID-19 infection are primarily due to respiratory failure, although a few people have died of multiple organ failure (MOF) or chronic comorbidities^4,5^. Therefore, reduced oxygen saturation is the major indicator of disease severity. However, symptoms at onset are relatively mild and a substantial proportion of patients have no obvious symptoms prior to the development of respiratory failure^4,5^. Clinically, it is difficult to stratify patients who may develop lethal COVID-19 until respiratory failure develops. Since early identification and effective treatment can reduce mortality and morbidity as well as relieve resource shortages, there is an urgent need for effective prediction models to identify patients who are most likely to develop respiratory failure and poor outcomes.

A robust, highly discriminatory and validated clinical prognosis model is required for stratification of these patients. Currently, very few reports propose reliable prediction models, which have been constructed using Transparent Reporting of a multivariable prediction model for Individual Prognosis Or Diagnosis (TRIPOD) guidance with internal and external validation^6^. One report based on a cohort of 191 COVID patients in early stages of the pandemic showed that age, Sequential Organ Failure Assessment (SOFA) score and D-dimer are independent risk factors of mortality^7^. However, D-dimer is not a routinely available clinical measurement and SOFA can be relatively low in COVID-19 patients on admission, but in the aforementioned report some patients in late stage of COVID-19 were transferred to its study hospitals^7^. This might explain the difference in SOFA scores.

In this study, we aimed to employ routine, widely available clinical test data to predict mortality of COVID-19 hospitalised patients using multivariable regression analysis in a cohort of 299 cases. The model has been established and validated with a new cohort of 145 patients with COVID-19 from a different hospital. Through this study, we hope that the experience from the outbreak in China will assist the rest of the world in better stratification of COVID-19 patients to achieve better outcomes for patients.

## Methods

### Study design and patients

This was a retrospective observational study performed in officially designated treatment centers for confirmed patients with COVID-19 in Wuhan at the center of the outbreak in China. The protocol was approved by the local Institutional Ethics Committee (Approval Number: KY-2020-10.02). First cohort of 299 patients admitted in Tongji hospital within January and February 2020 were enrolled into this study for model development. Second cohort of 151 patients admitted to Jinyintan hospital who were included in a previous study were reused for model validation. All patients were diagnosed by positive tests of novel coronavirus nucleic acids (SARS-CoV-2), according to WHO interim guidance^8^. Only patients who were discharged from hospital or had died were included in this study. Six patients in the second cohort, who died very quickly after admission and did not received any laboratory testing, were excluded. Patients younger than 18 years of age were also excluded.

### Data collection

Clinical data, including demographic information, chronic comorbidities, biomarkers of infection and other laboratory tests were collected from the local Online Medical System. The time from onset of symptoms to hospital admission was also recorded. SOFA was calculated within 24h of admission. For patients who did not have arterial blood gas analysis, peripheral capillary oxygen saturation (SpO_2_) to estimate the arterial oxygen partial pressure (PaO_2_) and the recorded fraction of inspired oxygen (FiO_2_) were used. The SOFA score for the respiratory system was calculated according to PaO_2_/FiO_2_ (P/F) ratio when patients received non-invasive or invasive mechanical ventilation. All patients were closely followed until they died or were discharged from the hospital. Hospital mortality and length of hospital stay were also recorded.

### Statistical analysis

Data were fully anonymized before data cleaning and analyses. We followed the TRIPOD guidance for development, validation and reporting of multivariable prediction models^6^. Analyses were performed in R version 3.5.1^9^. The development model was built using the data from Tongji Hospital. We performed logistic regression analysis with the outcome variable defined as mortality. Variables were selected *a priori* based on previous clinically-related studies, completeness across both sites, clinical knowledge and practicality of measurement in acute medical emergencies. Variables were excluded if they had high collinearity with global scores. The number of predictors was restricted based on the total number of outcomes in the development dataset^10^. Analysis was conducted for the following nine variables: age, hypertension, diabetes, SpO_2_, systolic blood pressure, albumin, lactate dehydrogenase (LDH), lymphocyte count and platelet count. Organ-specific markers, such as alanine transaminase (ALT) and blood urea nitrogen (BUN) were not included in the original development model because of collinearity with global markers, particularly LDH. SOFA score was not included for this reason, as explained in Results. D-dimer, interleukin (IL)-6 and cardiac troponin I (cTnI) were excluded from the analysis because they were not routinely tested in the cohorts. LDH was selected as a general marker for cell injury or death and since blood gas analysis were not commonly used in these hospitals, SpO_2_ was selected to reflect respiratory function and was modelled as a continuous variable to maximize statistical power^11^. Albumin was selected as a marker for the acute phase response instead of C reactive protein (CRP) because of CRP collinearity with LDH. Age is known risk factor for mortality from severe respiratory infections, including COVID-19. Lymphocyte count and platelet counts were reported in a previous publication of COVID-19 and selected^7,12^. In this study, lymphocyte count was log transformed due to extreme values. Systolic blood pressure (SBP), diabetes and hypertension were selected based on previous publications^7^. All nine selected variables were input into the multivariable model and backwards stepwise selection was performed with improvement in goodness-of-fit assessed by a reduction in the Akaike Information Criterion (AIC). A nomogram of the selected final model was used for graphical representation of the prediction model and was produced with R function regplot – this was chosen to offer maximum use of the prediction model in clinical practice without the need for internet access^13,14^.

Model performance was assessed via measures of discrimination and calibration. To assess discrimination, the C statistic was used. For calibration, patients in the development database were split into deciles, ordered by their probability of death. For each decile, the mean of the predicted probability of death was calculated and compared with the mean observed probability of death. Calibration plots were constructed including locally estimated scatterplot smoothing (loess) regression lines^15^.

Internal validity was assessed with bootstrapping (1000 replications) of the entire model building process including backwards stepwise selection of all potential predictors. Bootstrapped samples were created by drawing random samples with replacement from the development database. The prediction model was fitted on each bootstrap sample and tested on the original sample. Overfitting was assessed in each bootstrap replication. This allowed calculation of optimism and optimism adjusted discrimination and calibration statistics. The final development model coefficients were then adjusted for optimism using the optimism adjusted calibration slope for shrinkage^15^.

To assess external validity, the final optimism-adjusted model was applied to an independent data set, from Jinyintan hospital. External validity of the model was assessed using the C statistic, calibration-in-the-large, calibration slope and the calibration plot. Recalibration was conducted updating the intercept, and the intercept and slope.

## Results

### Patient cohorts

The development cohort included 299 patients who were admitted in Tongji Hospital. Data completeness is presented in supplementary Table 1. There were 155 deaths, the median age of patients was 65 years (IQR: 54-73) and 48·2% were male (Table 1). The validation cohort that included 145 patients was younger: age 56 years (IQR 47-68) with a higher percentage of males (65·5%) than in the developmental cohort (P<0·001). The median (IQR) time from onset of symptoms to hospital admission was no different in the developmental 10 [7-14] and validation 10 [7-13] cohorts. More patients had hypertension (42·6% vs 26·9%) and heart failure (4·4% vs 0.7%) in the development than validation cohorts, whilst diabetes (18·5% vs 11·7%), chronic obstructive pulmonary disease (COPD) (5·0% vs 3·4%) and others (6·4% vs 4·8) showed no statistical difference. SOFA scores were statistically higher in the development than validation cohorts (2·0 [2·0-4·0] vs 1·0 [0·0-3·0]) but both were very low compared to common critical illnesses. Table 2 shows that non-survivors were significantly older than survivors (69 years (IQR 61·75 to 75·00) vs. 52·50 (IQR 43·75-64·00), p<0·001). Patients with hypertension, showed a significantly higher mortality rate (p<0·001).

**Table 1:** Basic demographics and clinical features of patients in the developmental and validation datasets. (For continuous variables median and IQR are reported)

|  | Developmental | Validation |  |  |
| --- | --- | --- | --- | --- |
|  | Tongji (N=299) | Jinyintan (N=145) | Total (N=444) | P value |
| Age (Years) | 65·0 (54·0, 73·0) | 56·0 (47·0, 68·0) | 62·0 (50·8, 71·0) | < 0·001 |
| Gender |  |  |  | < 0·001 |
| Male | 144 (48·2%) | 95 (65·5%) | 239 (53·8%) |  |
| Female | 155 (51·8%) | 50 (34·5%) | 205 (46·2%) |  |
| Days since onset | 10·0 (7·0, 14·0) | 10·0 (7·0, 13·0) | 10·0 (7·0, 14·0) | 0·357 |
| Co-morbidities |  |  |  |  |
| Hypertension | 127 (42·6%) | 40 (27·6%) | 167 (37·6%) | 0·001 |
| Diabetes | 55 (18·5%) | 17 (11·7%) | 72 (16·3%) | 0·069 |
| COPD | 15 (5·0%) | 5 (3·4%) | 20 (4·5%) | 0·627 |
| Heart Failure | 13 (4·4%) | 2 (1·4%) | 15 (3·4%) | 0·042 |
| Smoking | 10 (3·4%) | 7 (4·8%) | 17 (3·8%) | 0·449 |
| Other | 19 (6·4%) | 7 (4·8%) | 26 (5·9%) | 0·515 |
| Mortality, n (%) | 155 (51·8%) | 69 (47·6%) | 224 (50·5%) | 0·419 |
| SPO<sub>2</sub>, (%) | 95·0 (90·0, 98·0) | 95·0 (89·0, 97·0) | 95·0 (90·0, 98·0) | 0·366 |
| SPO<sub>2</sub>, (%) |  |  |  | 0·340 |
| ≥90 | 229 (77·4%) | 95 (73·1%) | 324 (76·1%) |  |
| <90 | 67 (22·6%) | 35 (26·9%) | 102 (23·9%) |  |
| SOFA score | 2·0 (2·0, 4·0) | 1·0 (0·0, 3·0) | 2·0 (1·0, 4·0) | < 0·001 |
| Systolic blood pressure, mmHg | 132·5 (118·0, 145·0) | 122·5 (117·0, 137·3) | 129·5 (117·0, 143·0) | 0·008 |
| WBC, 10<sup>9</sup>/L | 6·8 (4·7, 10·6) | 7·6 (5·0, 11·2) | 6·9 (4·8, 10·8) | 0·562 |
| Lymphocyte count, 10<sup>9</sup>/L | 0·75 (0·50, 1·11) | 0·74 (0·52, 1·13) | 0·75 (0·50, 1·12) | 0·703 |
| Platelet count, 10<sup>9</sup>/L | 179·5 (137·3, 247·8) | 192·0 (140·5, 242·3) | 182·0 (138·0, 247·3) | 0·413 |
| D-dimer, mg/L |  |  |  | 0·274 |
| <0·5 | 49 (19·0%) | 36 (25·9%) | 85 (21·4%) |  |
| 0·5-1·0 | 55 (21·3%) | 28 (20·1%) | 83 (20·9%) |  |
| ≥1·0 | 154 (59·7%) | 75 (54·0%) | 229 (57·7%) |  |
| Albumin, g/L | 33·4 (29·9, 36·2) | 30·7 (27·4, 34·5) | 32·5 (28·9, 35·8) | < 0·001 |
| hs-cTnI, pg/ml |  |  |  | 0·198 |
| <28·0 | 173 (71·5%) | 107 (77·5%) | 280 (73·7%) |  |
| ≥28·0 | 69 (28·5%) | 31 (22·5%) | 100 (26·3%) |  |
| ALT, U/L | 26·0 (16·0, 40·8) | 29·0 (19·0, 51·0) | 27·0 (17·0, 45·0) | 0·017 |
| AST, U/L | 33·5 (24·0, 56·0) | 36·0 (27·0, 47·5) | 34·0 (25·0, 53·0) | 0·400 |
| Bilirubin, μmol/L | 10·3 (7·6, 14·6) | 12·5 (9·7, 17·4) | 11·4 (8·2, 15·9) | <0·001· |
| BUN, mmol/L | 5·9 (3·9, 9·2) | 5·8 (4·4, 7·6) | 5·9 (4·1, 8·6) | 0·602 |
| Creatinine, U/L | 74·0 (58·0, 95·0) | 72·9 (58·8, 84·6) | 73·2 (58·5, 93·0) | 0·264 |
| Creatinine kinase, U/L | 131·0 (70·0, 380·8) | 92·0 (52·0, 183·0) | 101·0 (58·0, 238·0) | 0·005 |
| LDH, U/L | 384.0 (264.0, 541.0) | 333.0 (251.0, 521.0) | 362.0 (260.5, 535.3) | 0.199 |
| CRP, mg/dl | 65.3 (27.4, 113.9) | 58.3 (23.7, 114.5) | 64.5 (25.1, 114.4) | 0.442 |
| INR, s | 1.13 (1.04, 1.25) | 0.96 (0.90, 1.03) | 1.07 (0.98, 1.18) | <0.001 |
| Procalcitonin, ng/ml | 0.13 (0.05, 0.37) | 0.07 (0.05, 0.19) | 0.10 (0.05, 0.27) | 0.030 |

Data are presented as median (IQR), or n (%). P values were calculated using Wilcoxon rank-sum test, $\chi^2$ or Fisher's exact test. ALT= Alanine aminotransferase. AST= Aspartate aminotransferase. BUN=Blood urea nitrogen. COPD=chronic obstructive disease. CRP= C-reactive protein. hs-cTnI= high-sensitivity cardiac troponin I. INR= international normalized ratio. LDH= lactate dehydrogenase. SOFA= sequential organ failure assessment. SPO<sub>2</sub>= pulse oxygen saturation. WBC=white blood cell.

**Table 2:** Differences in demographics between COVID-19 patients who survived and died in developmental and validation datasets. (For continuous variables median and IQR are reported)

|  | Developmental (Tongji) |  |  | Validation (Jinyintan) |  |  |
| --- | --- | --- | --- | --- | --- | --- |
|  | Survived (N=144) | Died (N=155) | P value | Survived (N=76) | Died (N=69) | P value |
| Age (Years) | 56.0 (47.8, 67.0) | 69.00 (62.0, 76.0) | <0.001 | 47.0 (40.8, 53.3) | 67.0 (61.0, 73.0) | < 0.001 |
| Gender |  |  | 0.191 |  |  | 0.673 |
| Male | 75 (52.1%) | 69 (44.5%) |  | 51 (67.1%) | 44 (63.8%) |  |
| Female | 69 (47.9%) | 86 (55.5%) |  | 25 (32.9%) | 25 (36.2%) |  |
| Days since onset | 10.0 (7.0, 13.0) | 10.0 (7.0, 15.0) | 0.267 | 9.0 (7.0, 12.3) | 10.0 (7.0, 13.0) | 0.062 |
| Co-morbidities |  |  |  |  |  |  |
| Hypertension | 47 (32.6%) | 80 (51.9%) | <0.001 | 8 (10.5%) | 31 (44.9%) | <0.001 |
| Diabetes | 21 (14.6%) | 34 (22.2%) | 0.09 | 4 (5.3%) | 13 (18.8%) | 0.018 |
| COPD | 2 (1.4%) | 13 (8.4%) | 0.006 | 1 (1.3%) | 4 (5.8%) | 0.192 |
| Heart Failure | 1 (0.7%) | 12 (7.8%) | 0.003 | 0 (0.0%) | 1 (1.4%) | 0.476 |
| Smoking | 5 (3.5%) | 5 (3.2%) | 0.914 | 1 (1.3%) | 6 (8.7%) | 0.054 |
| Other | 8 (5.6%) | 11 (7.1%) | 0.575 | 2 (2.6%) | 5 (7.2%) | 0.258 |
| SpO<sub>2</sub>, (%) | 97.0 (95.0, 98.0) | 92.0 (83.8, 96.0) | <0.001 | 97.0 (94.8, 98.0) | 89.0 (81.0, 94.0) | < 0.001 |
| SpO<sub>2</sub>, (%) |  |  | <0.001 |  |  | < 0.001 |
| ≥90 | 136 (94.4%) | 93 (61.29%) |  | 68 (94.4%) | 27 (46.6%) |  |
| <90 | 8 (5.6%) | 59 (38.8%) |  | 4 (5.6%) | 31 (53.4%) |  |
| SOFA score | 2.0 (1.0, 2.0) | 4.0 (2.0, 5.0) | <0.001 | 1.0 (0.0, 1.0) | 3.0(2.0, 5.0) | < 0.001 |
| Systolic blood pressure, mmHg | 131.0 (120.0, 141.3) | 133.5 (116.3, 149.0) | 0.169 | 119.0 (115.0, 125.3) | 131.5 (122.0, 145.3) | < 0.001 |
| WBC, 10<sup>9</sup>/L | 5.4 (4.0, 7.2) | 9.0 (5.9, 13.4) | <0.001 | 5.6 (3.5, 8.8) | 8.7 (6.7, 12.5) | < 0.001 |
| Lymphocyte count, 10<sup>9</sup>/L | 0.98 (0.75, 1.52) | 0.57 (0.43, 0.82) | <0.001 | 1.00 (0.74, 1.31) | 0.54 (0.39, 0.73) | <0.001 |
| Platelet count, 10<sup>9</sup>/L | 207.0 (161.0, 278.0) | 159.0 (114.5, 220.5) | <0.001 | 194.0 (147.5, 274.5) | 188.0 (130.0, 228.5) | 0.065 |
| D-dimer, mg/L |  |  | <0.001 |  |  | <0.001 |
| <0.5 | 39 (32.0%) | 10 (7.4%) |  | 33 (45.2%) | 3 (4.5%) |  |
| 0.5-1.0 | 37 (30.3%) | 18 (13.2%) |  | 18 (24.7%) | 10 (15.2%) |  |
| ≥ 1.0 | 46 (37.7%) | 108 (79.4%) |  | 22 (30.1%) | 53 (80.3%) |  |
| Albumin g/L | 35.6 (32.7, 37.9) | 31.3 (28.2, 34.3) | <0.001 | 33.8 (30.7, 35.7) | 28.5 (25.8, 30.4) | <0.001 |
| cTnI, pg/ml |  |  | <0.001 |  |  | <0.001 |
| <28.0 | 104 (96.3%) | 69 (51.5%) |  | 70 (98.6%) | 37 (55.2%) |  |
| ≥28.0 | 4 (3.7%) | 65 (48.5%) |  | 1 (1.4%) | 30 (44.8%) |  |
| ALT, U/L | 23.0 (15.0, 37.0) | 28.0 (19.0, 44.0) | 0.026 | 26.5 (18.0, 40.3) | 41.0 (20.5, 55.5) | 0.024 |
| AST, U/L | 30.0 (21.5, 44.0) | 42.0 (26.0, 64.0) | <0.001 | 31.0 (25.8, 39.0) | 42.0 (34.5, 59.5) | <0.001 |
| Bilirubin, μmol/L | 8.4 (6.5, 11.9) | 12.7 (9.0, 18.8) | <0.001 | 11.5 (9.1, 13.8) | 15.7 (11.6, 22.6) | <0.001 |
| BUN, mmol/L | 4.4 (3.3, 6.0) | 8.0 (5.7, 11.9) | <0.001 | 4.6 (3.7, 5.8) | 7.5 (6.2, 8.9) | <0.001 |
| Creatinine, U/L | 66.0 (56.0, 83.0) | 85.0 (66.0, 109.0) | <0.001 | 71.8 (57.6, 82.1) | 74.4 (64.6, 92.1) | 0.180 |
| Creatinine kinase, U/L | 90.5 (38.9, 383.8) | 137.0 (78.0, 381) | 0.052 | 76.5 (46.8, 144.3) | 99.0 (58.0, 196) | 0.052 |
| LDH, U/L | 295 (216, 388) | 505(371, 676) | <0.001 | 264 (214, 319) | 531 (413, 637) | <0.001 |
| CRP, mg/dl | 39.0 (10.7, 80.6) | 97.2 (47.2, 148.8) | <0.001 | 26.4 (7.5, 61.8) | 112.1 (65.4, 160) | <0.001 |
| INR, s | 1.08 (1.02, 1.16) | 1.20 (1.08, 1.38) | <0.001 | 0.92 (0.86, 0.99) | 1.01 (0.94, 1.10) | <0.001 |

Data are presented as median (IQR), or n (%). P values were calculated using Wilcoxon rank-sum test, $\chi^2$ or Fisher's exact test. ALT= Alanine aminotransferase. AST= Aspartate aminotransferase. BUN=Blood urea nitrogen. COPD=chronic obstructive disease. CRP= C-reactive protein. hs-cTnI= high-sensitivity cardiac troponin I. INR= international normalized ratio. LDH= lactate dehydrogenase. SOFA= sequential organ failure assessment. SPO<sub>2</sub>= pulse oxygen saturation. WBC=white blood cell.

### Model development

In the developmental cohort (Tongji Hospital), 268/299 patients had complete data for all nine variables. The final multivariable model included age (adjusted OR 1·054; 95% CI 1·028 to 1·083), LDH (adjusted OR 1·004; 95% CI 1·002 to 1·006), log lymphocyte count (adjusted OR 0·296; 95% CI 0·148 to 0·541) and SpO_2_(adjusted OR 0·897; 95% CI 0·831 to 0·958) (Table 3). The final model showed excellent discrimination (C statistic = 0·89) and was well calibrated (slope =1 and calibration-in-the-large =0·00), as shown in the calibration plot (Table 4 and Figure 1). The final model for mortality is presented as a nomogram and an example of how to use the nomogram is presented for a patient aged 59, with LDH of 482, SpO_2_ of 85% and lymphocyte count of 0·64 (Figure 2).

**Table 3:**
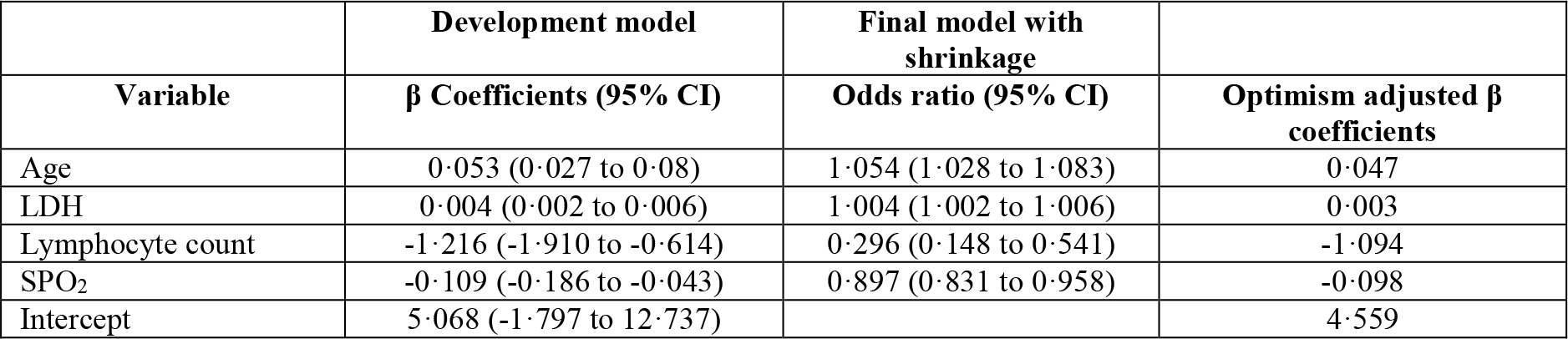
Final multivariable model in development dataset and optimism adjusted β coefficients.

|  | Development model | Final model with shrinkage |  |
| --- | --- | --- | --- |
| Variable | $\beta$ Coefficients (95% CI) | Odds ratio (95% CI) | Optimism adjusted $\beta$ coefficients |
| Age | 0.053 (0.027 to 0.08) | 1.054 (1.028 to 1.083) | 0.047 |
| LDH | 0.004 (0.002 to 0.006) | 1.004 (1.002 to 1.006) | 0.003 |
| Lymphocyte count | -1.216 (-1.910 to -0.614) | 0.296 (0.148 to 0.541) | -1.094 |
| SPO<sub>2</sub> | -0.109 (-0.186 to -0.043) | 0.897 (0.831 to 0.958) | -0.098 |
| Intercept | 5.068 (-1.797 to 12.737) |  | 4.559 |

**Table 4.** Internal validation: model performance.

| Measure | Development model | Average optimism | Optimism adjusted |
| --- | --- | --- | --- |
| C statistic | 0.893 (0.856 to 0.930) | 0.013 | 0.880 |
| Calibration in the large | 0.000 (-0.331 to 0.330) | -0.006 | 0.006 |
| Calibration Slope | 1.000 (0.773 to 1.264)) | 0.100 | 0.900 |

**Figure 1.**
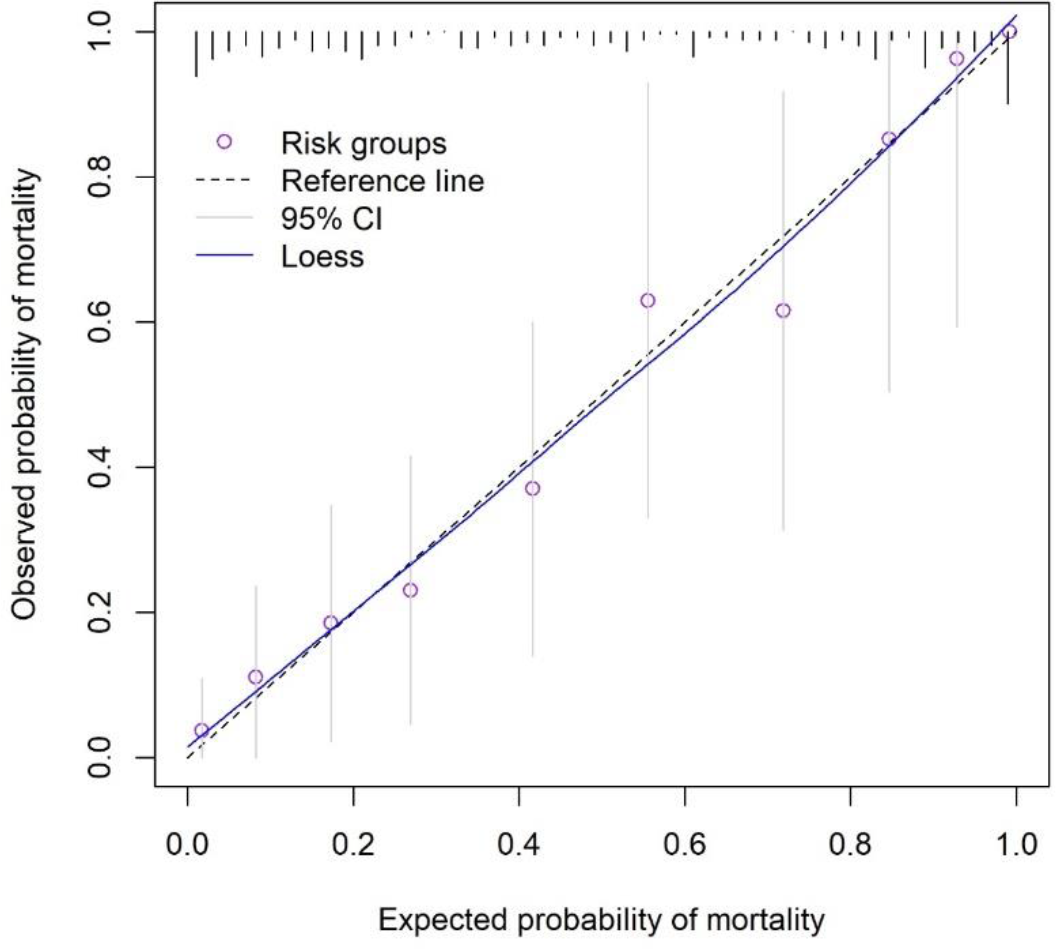
Calibration plot of the final development model.

**Figure 2.**
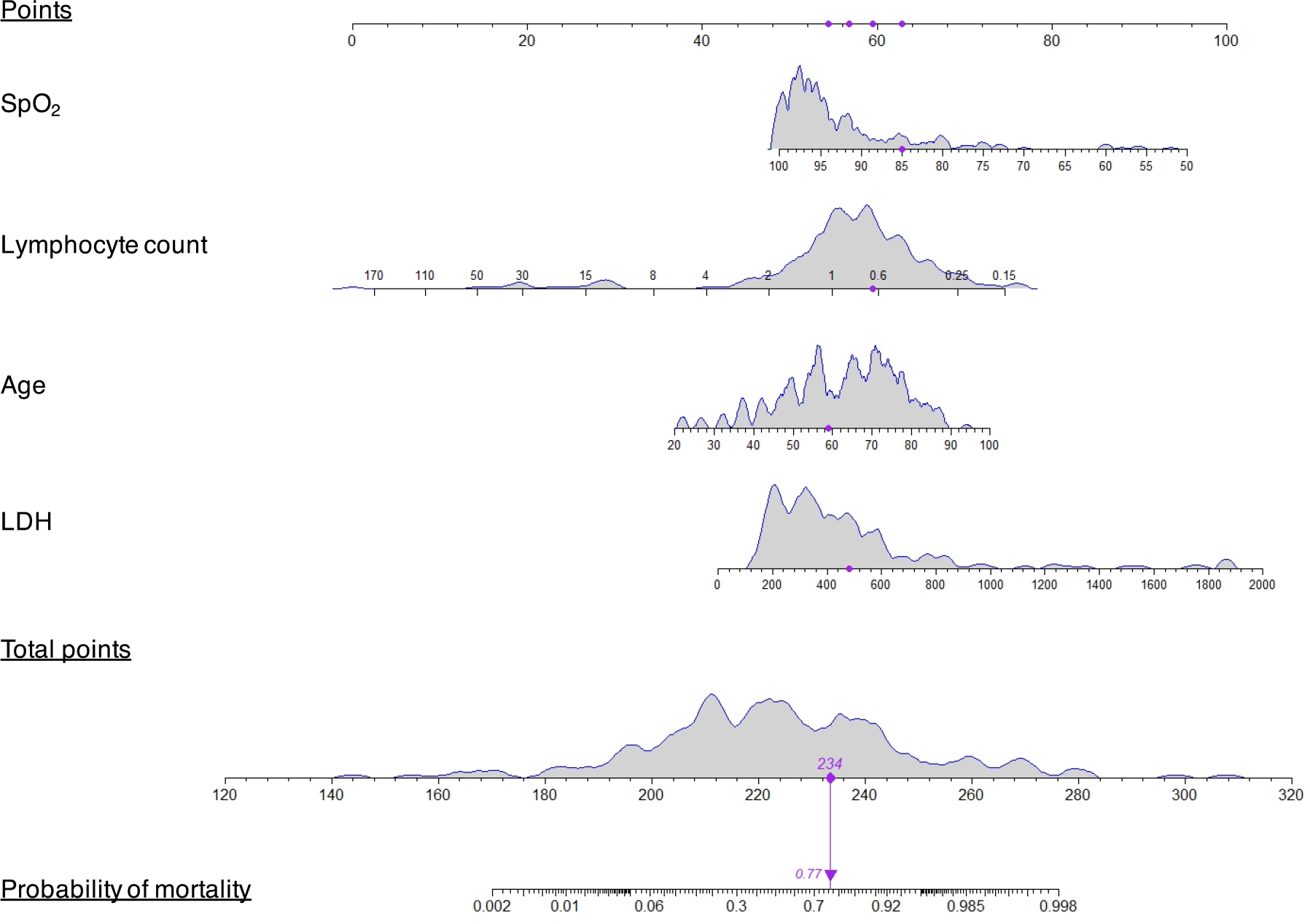
Nomogram of the final development model. The nomogram presents the distribution of continuous variables and for categorical variables the relative contribution of variable to the model is reflected in the size. The distribution of the final score is shown for the development data and the red dots represent a randomly selected patient from the development dataset with probability of mortality of 0.77.

### Internal validation

Interval validation was run using bootstrap resampling and showed a small amount of overfitting. Bootstrapping with backwards stepwise selection provided a shrinkage factor of 0.900. After adjusting for overfitting the final model retained high discrimination (C statistic= 0·880) and calibration-in-the-large of 0·006 (Table 4). The shrinkage factor was applied to the β coefficients of the multivariable model to provide optimism adjusted coefficients (Table 3).

### External validation

External validation is required because the accuracy of a predictive model will always be high if it is validated on the development cohort used to generate the model^16,17^. In this study, we applied our final prediction model with optimism adjusted coefficients to the validation dataset. There were 145 patients in the validation dataset of whom 69 patients (47·6%) died. Of the 145 patients, 127 had complete data for the four variables in the final developmental model (age, lymphocyte count, SpO_2_ and LDH) The C statistic for the discrimination of the developed model in the independent data was 0·980 (0·958 to 1·000) and the model had reasonable calibration (slope and calibration-in-the-large) (Table 5). Figure 3 suggests some mis-calibration of the model in the independent data with over-estimation of risk of mortality in low-risk individuals and under-estimation of risk in high-risk individuals. After recalibration of the intercept, the discrimination of the model was the same (as expected given that recalibration does not influence the ranking of individuals according to the model) as was the calibration slope. Calibration-in-the-large was 0·00 (−0·477 to 0·471). The calibration plots still suggest mis-calibration of the model (Figure 3c). When both intercept and slope were recalibrated the calibration slope was 1·00 (0·675 to 1·477) and the plot then demonstrated good calibration (Figure 3d).

**Table 5.** External validation and recalibration: model performance.

| Measure | Validation without shrinkage | Validation with shrinkage | Recalibration (intercept only) | Recalibration (intercept and slope) |
| --- | --- | --- | --- | --- |
| C statistic | 0.980 (0.958 to 1.000) | 0.980 (0.958 to 1.000) | 0.980 (0.958 to 1.000) | 0.980 (0.958 to 1.000) |
| Calibration in the large | -0.192 (-0.690 to 0.300) | -0.195 (-0.673 to 0.276) | 0.000 (-0.477 to 0.471) | 0.000 (-0.725 to 0.738) |
| Calibration Slope | 2.227 (1.503 to 3.291) | 2.476 (1.671 to 3.658) | 2.476 (1.671 to 3.658) | 1.00 (0.675 to 1.477) |

**Figure 3.**
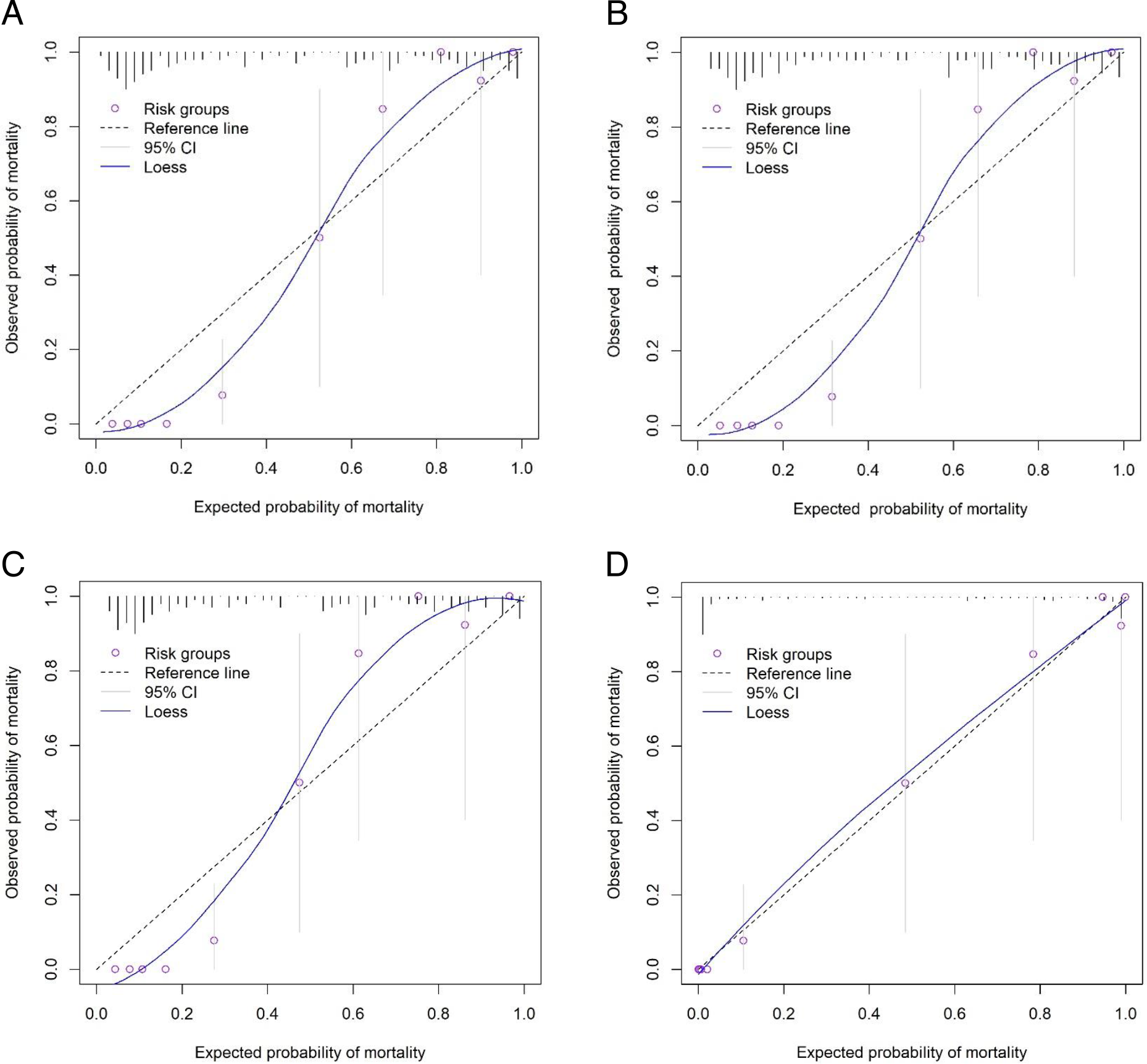
Calibration plots of external validation. A) Calibration without shrinkage B) Calibration with shrinkage of final model coefficients. C) Recalibration with intercept updated. D) Recalibration with slope and intercept updated.

## Discussion

Most routine scoring systems, such as SOFA, which are very useful in the intensive care unit (ICU) for sepsis and other critical illnesses^18-20^, were very low in COVID-19 patients on admission with median SOFA score of 2. However, 155/299 and 69/145 patients in the development and validation cohorts died in the hospital, respectively. This poor outcome strongly indicates that these routine scoring systems used in general wards and ICU cannot accurately assess the severity and predict the mortality of patients with COVID-19. We have therefore used a hospitalized study population from Wuhan Tongji Hospital to develop a clinical prediction model using available demographic and clinical indicators. The most informative factors in COVID-19 positive patients were age, LDH, lymphocyte count and SpO_2_. The developed statistical model had good discrimination and minimal model optimism. In external validation using an independent cohort from Jinyintan hospital, the model had excellent discrimination but needed recalibrating to account for the different underlying risk profile in the independent dataset. Specifically, the development and validation cohorts are temporally distinct as the validation cohort were from earlier in the COVID-19 outbreak. In addition, they are from different sites within Wuhan and the demographics are distinct, with patients admitted to the Jinyintan hospital being younger with a male majority and less patients with hypertension and low SOFA scores (1.0 [0-3] vs 2.0 [2.0-4.0]. However, the nomogram of the final model after optimization can be used by clinicians to give a prediction of mortality and thus inform treatment choice and guide patient and family counselling.

Old age is a major factor of mortality from infection^21^. This is consistent with the current data from China on COVID-19 with less than 1% in children <10 years old, increasing to 2%, 4% and 10% in age 11-40, 41-70, and >70, respectively. However, the majority of people infected are 30-70 years old (approximately 80% of the total cases), thereby making it difficult to stratify the majority of cases according to age. Patient comorbidities, including hypertension, diabetes, did not significantly contribute to the model, although mortality rates were significantly higher in patients with hypertension.

The pathological feature of COVID-19 is mainly that of a viral pneumonia with alveolar oedema and blockage of small bronchi to compromise gas exchange within the lungs. Therefore, PaO_2_ will drop when severe pneumonia develops. Since arterial blood gas (ABG) analysis is an invasive and complex process, which requires arterial blood taking and use of a ABG analyzer, SpO_2_ measurement is much more used for continuous analysis of blood oxygen saturation in patients to estimate the arterial oxygen partial pressure (PaO_2_)^22^. Moreover, many mobile phone brands contain built-in sensors and SpO_2_ can be measured by patients themselves. In our cohorts, many patients had no blood gas data and therefore SpO_2_ was used as a variable in our model. Although SpO_2_ is not as accurate as PaO_2_ in critical illnesses, it could give the first indication of compromised lung gas exchange in the early stages of COVID-19, which would then be verified by ABG measurement upon admission to hospital.

Lymphopenia is very common in patients with influenza virus infection and bacterial infection, particularly in sepsis^23,24^. CD4 and CD8 T cell apoptosis causes prolonged infection by promoting virus survival^25^, and induces immune suppression to increase mortality in patients with sepsis^26^. In this study, we found a dramatic difference in lymphocyte counts between survivors and non-survivors. In non-survivors, the peripheral lymphocyte counts were very low on admission, indicating that extensive lymphocyte death occurred following infection. LDH is released into circulation after cell injury or death^27^ and serves as an index of the extent of cellular damage by the virus or the host immune response^28^.

This study has several limitations. At this stage in the global outbreak of COVID-19, no published studies have developed and validated prediction models for assessing the probability of mortality in hospitalized patients. A most recent report proposed SOFA score, age and D-dimers as the important prognostic markers^7^. However, in our cohorts, the SOFA score was very low although there is statistical difference between survivors and non-survivors. This difference may be due to some patients being treated in other hospitals prior to transfer in the previously reported study^7^. In addition, SOFA score relies on blood gas analysis, which cannot be easily done continually, in clinics and in under-resourced settings, thus making it difficult for clinicians to use readily. In addition, since D-dimer is not routinely requested, too many patients in our cohorts had no test performed, making it is difficult for us to verify published results. Organ-specific injury markers were not included in the model, including cTnI due to missing data (not routinely measured), as well as ALT and BUN due to high co-linearity with other variables within the model. Therefore, the choice of potential prognostic factors could not be informed by a systematic review of existing prediction models. Similarly, there are no published estimates of discrimination and calibration to compare this study to. Additionally, the event rate is limited, especially in the independent dataset where at least 100 events would be preferable^29^. A multicenter study with a larger cohort for model development and also for validation would add more power, reduce bias, and make the findings more generically applicable.

There is also inherent bias in the recruitment of patients because the Jinyintan hospital was the first officially designated receiving hospital in Wuhan for all COVID-19 cases at the start of the outbreak. The recruited patients were mainly self-referrals, but some were referred by other hospitals immediately after a positive diagnostic test. Self-referred patients are more likely to have severe symptoms that cause them to seek emergency medical help. Likewise, other hospitals are more likely to test patients who have more pronounced symptoms, thus creating an inherent bias in the patient sample. Therefore, the cohort may not accurately represent patients with mild or asymptomatic COVID-19. These results need to be further validated with patients who are hospitalized for different severities of illness.

Despite these limitations, the obvious advantage of the model developed here is that the panel of variables used to build the model are basic demographics and values from accessible clinical tests that are recorded routinely in the acute clinical setting. With the vivid nomogram, this model will enable the majority of clinicians to assess the severity of patient illness without difficulty, particular in very busy clinical settings. We were also able to externally validate our model in an independent dataset. The discrimination of the model was high in both internal and external validation showing that the model is able to accurately differentiate high and low risk individuals. The prediction models are based on and validated in Chinese hospitalized COVID-19 positive populations and should therefore be applicable to other sites within China and may also be translatable into other sites during the global outbreak. However, separate validation will be required in other patient populations before widespread application.

In conclusion, traditional early warning scores and organ injury markers do not help to predict severity and mortality in patients with COVID-19. Here, we have developed and validated a clinical prediction model using readily available clinical markers which is able to accurately predict mortality in people with COVID-19. This model can therefore be used in current pandemic regions to identify patients who are at risk of severe disease at an early enough stage to initialize intensive supportive treatment and to guide discussions between clinicians, patients and families. It has the potential to be used throughout the world following additional validation in relevant worldwide datasets, with recalibration as necessary to account for differing risk profiles of patients.

## Data Availability

Aggregated data will be made available on request.

## Acknowledgements

This study was supported by following funding: Key Research and Development Plan of Jiangsu Province (BE2018743 and BE2019749), National Institute for Health Research (NIHR) (PDF-2018-11-ST2-006), British Heart Foundation (BHF) (PG/16/65/32313) and Liverpool University Hospitals NHS Foundation Trust in UK. We also thank all the patients and medical staff involved.

## Author Contribution

JX, HC, SL, GW, BD and HQ had the idea for and designed the study; JX, HC, SL, YW, HK, RZ, XL and ZT collected the clinical data. DH, STA and LB did the statistical analysis and wrote part of the manuscript. GW and CHT wrote the manuscript. All authors contributed to acquisition, analysis, or interpretation of data. All authors revised the report and approved the final version before submission.

